# Large-scale silane bead-based SARS-CoV-2 testing of a nursing home in Spain identifies a viral reservoir during lockdown period

**DOI:** 10.1101/2021.02.08.21251358

**Authors:** Nicholas D. Weber, Ainhoa Goñi-Salaverri, Jose A. Rodríguez, Juan Pablo Unfried, Daniel Alameda, Mirian Fernández-Alonso, Elena Sáez, Sheila Maestro-Galilea, Felix Alegre, Francisco Carmona-Torre, Marta Marin-Oto, Cristina Olagüe, Leticia Odriozola, Mar Navarro-Alonso, Rafael Sanchez-Ostiz, Josepmaria Argemi, Jose Luis del Pozo, David Lara-Astiaso

**Affiliations:** Vivet Therapeutics SL, Pamplona, Spain; Oncohematology Department, Cima Universidad de Navarra, Pamplona, Spain; Genomics Unit, Cima Universidad de Navarra, Pamplona, Spain; Department of Gene Therapy and Regulation of Gene Expression, Cima Universidad de Navarra, Pamplona, Spain; Microbiology Unit. Clínica Universidad de Navarra, Pamplona, Spain; Medicinal Chemistry Laboratory, Department of Molecular Therapy, CIMA Universidad de Navarra, Pamplona, Spain; Internal Medicine Department, Clínica Universidad de Navarra, Pamplona, Spain; Infectious Diseases Division and Clinical Microbiology, Clínica Universidad de Navarra, Pamplona, Spain; Respiratory Medicine Department, Clínica Universidad de Navarra, Pamplona, Spain; Idea Innovacion, Cizur Menor, Navarra, Spain; Liver Unit, Internal Medicine Department. Clínica Universidad de Navarra, Pamplona, Spain; Division of Medicine - Gastroenterology and Hepatology Department, University of Pittsburgh. Pittsburgh, PA USA; Wellcome-MRC Cambridge Stem Cell Institute, Cambridge, England

## Abstract

**Background:** Spain is one of the countries most heavily affected by the COVID-19 pandemic. As in other countries such as UK and USA, nursing homes have been an important human reservoir for the virus and the population with the highest mortality worldwide. The presence of asymptomatic carriers within nursing homes is one of the factors that could provoke new outbreaks during the relaxing of lockdown measures.

**Methods:** We developed a high-throughput protocol for RNA extraction of patient samples based on silane magnetic beads in multi-well plates. The sensitivity, specificity and reproducibility rates were assessed using positive and negative clinical samples from the Clinica Universidad de Navarra, Spain. We utilized the protocol to test a pilot cohort of 138 residents and 87 staff from a nursing home in Northern Navarre, Spain.

**Findings:** Our protocol showed high sensitivity (100%), specificity (96·0%) and linear correlation with PCR cycle threshold values obtained with a standard testing kit (R^2^ = 0·807, p=3E-05). Testing of 225 individuals from the nursing home revealed 63 residents (46%) and 14 staff (16%) positive for SARS-CoV-2. Only 18 of the positive residents (28·6%) were symptomatic at time of testing. During follow-up, 6 PCR-negative symptomatic residents were retested and resulted positive. One-month mortality among positive residents was higher than in negative residents (15·9% vs 1·3%), regardless of age or comorbidities.

**Interpretation:** Rapid silane bead-based RNA extraction expanded the testing capabilities and COVID-19 patients were promptly identified. Personal and public health measures were enacted to avoid spreading and tighten clinical surveillance. The ability to easily adapt the technical capabilities of academic research centers to large-scale testing for SARS-CoV-2 could provide an invaluable tool for ensuring a safe lifting of lockdown in countries with high numbers of cases.

**Funding:** European Molecular Biology Organization and Genomics Unit, Cima Universidad de Navarra.

## Introduction

Spain has been one of the most severely affected countries by the COVID-19 pandemic, with the second highest fatality rate worldwide.^1,2^ Recent regional data estimates have shown that 17,585 residents of nursing homes in Spain could have died from COVID-19. Since many of these people were not tested, the mortality rate above global COVID-19 figures is unknown.^3^ Although physical distancing and lockdown of the entire population have decreased the spread of SARS-CoV-2 and daily mortality, nursing homes have been an important focus of contagion during the pandemic period, probably due to difficulties in providing reliable individual isolation, and presymptomatic or asymptomatic transmission rates.^4^ While there is an urgent need to lift lockdown measures, specific strategies have to be in place to avoid *de novo* infections from viral carriers that could lead to new outbreaks in the next weeks or months due to the high prevalence of non-protected subjects.^5,6^ PCR testing is the only way to detect viral carriers among people at risk.^7,8^ However, the development of rapid large-scale SARS-CoV-2 PCR strategies has been hampered by the need for commercial RNA extraction kits and intensive manual labor,^9,10^ which increases the time and costs of these analyses and limits their overall throughput. As reported by several media outlets in UK and USA, research laboratories are a valuable untapped resource for providing PCR testing capabilities.^8,11^ Here we show a novel labor-, cost-, and time-saving protocol of RNA extraction, using silane beads (SB) and magnetized pelleting followed by one-step RT-qPCR (Figure 1). With a case study of a nursing home in Northern Navarre, Spain, we show that this method is suitable for sensitive and timely detection of SARS-CoV-2 in vulnerable communities and health care workers. The entire protocol from the receipt of patient samples to the analysis of data can be performed by a team of four workers processing 1152 samples in a typical workday.

**Figure 1.**
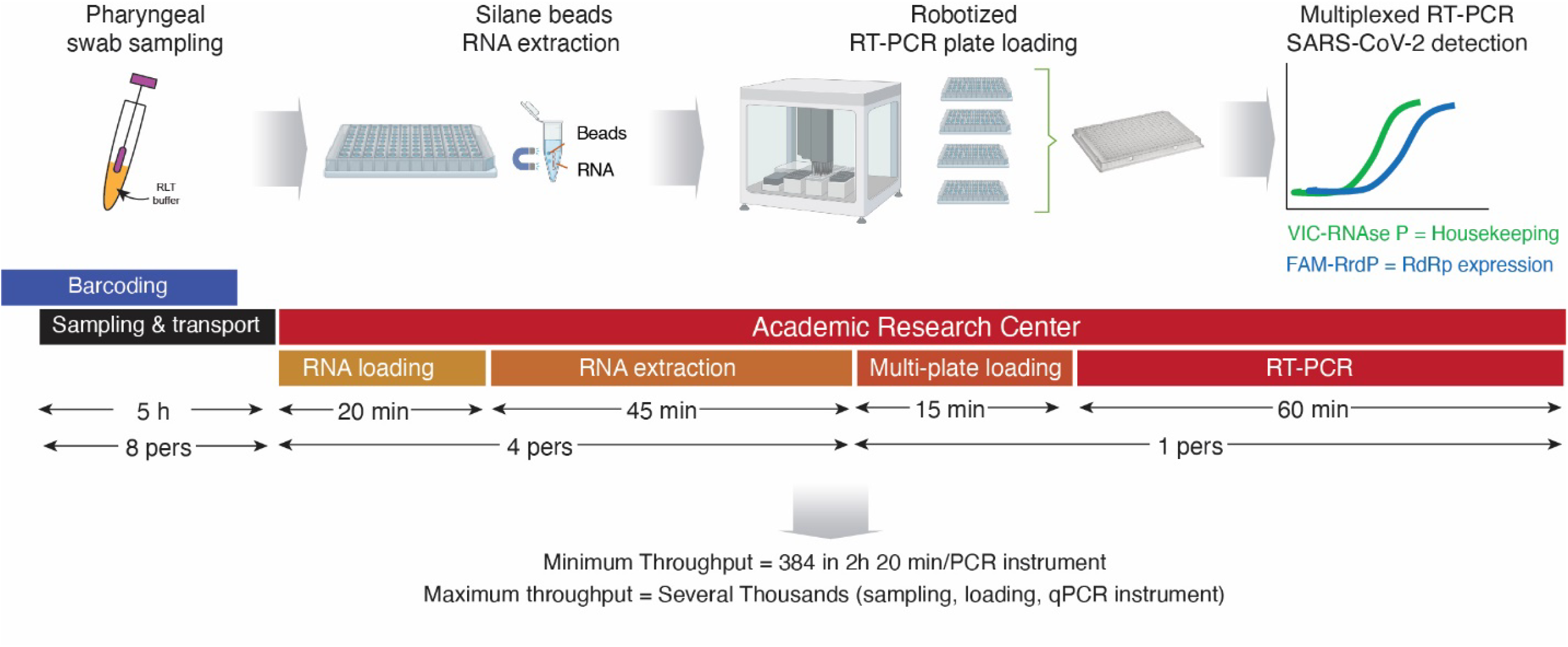
Schematic Representation of silane bead-based large-scale RNA extraction and RT-qPCR. After barcoding of collection tubes, the individuals are sampled using oropharyngeal swabs by trained personnel with adequate personal protection equipment. Each tube contains 800 μL of Buffer RLT Plus that inactivates viral particles. Once the tubes reach the laboratory in the academical research center, 150 μL of sample in buffer are loaded in a 96 well plate. RNA is extracted in the 96-well plate via silane magnetic beads and multichannel pipetting. The eluted RNA is loaded via a robot onto a 96- or 384-well one-step RT-qPCR plate, where it undergoes thermocycling and target DNA detection.

## Results

We first compared two magnetic bead-based RNA extraction protocols (SB vs Oligo-dT Dynabeads) using 16 patients’ deidentified nasopharyngeal swabs from SARS-CoV-2-positive and negative patients. Swab samples were transferred to 96-well plates and extracted with either method utilizing magnetized pelleting. Then, single-step reverse transcription quantitative polymerase chain reaction (RT-qPCR) for SARS-CoV-2 detection was performed. SB extraction resulted in a more sensitive detection of both SARS-CoV-2 (RdRp) and thus, was selected for further testing (Figure 2A).

**Figure 2.**
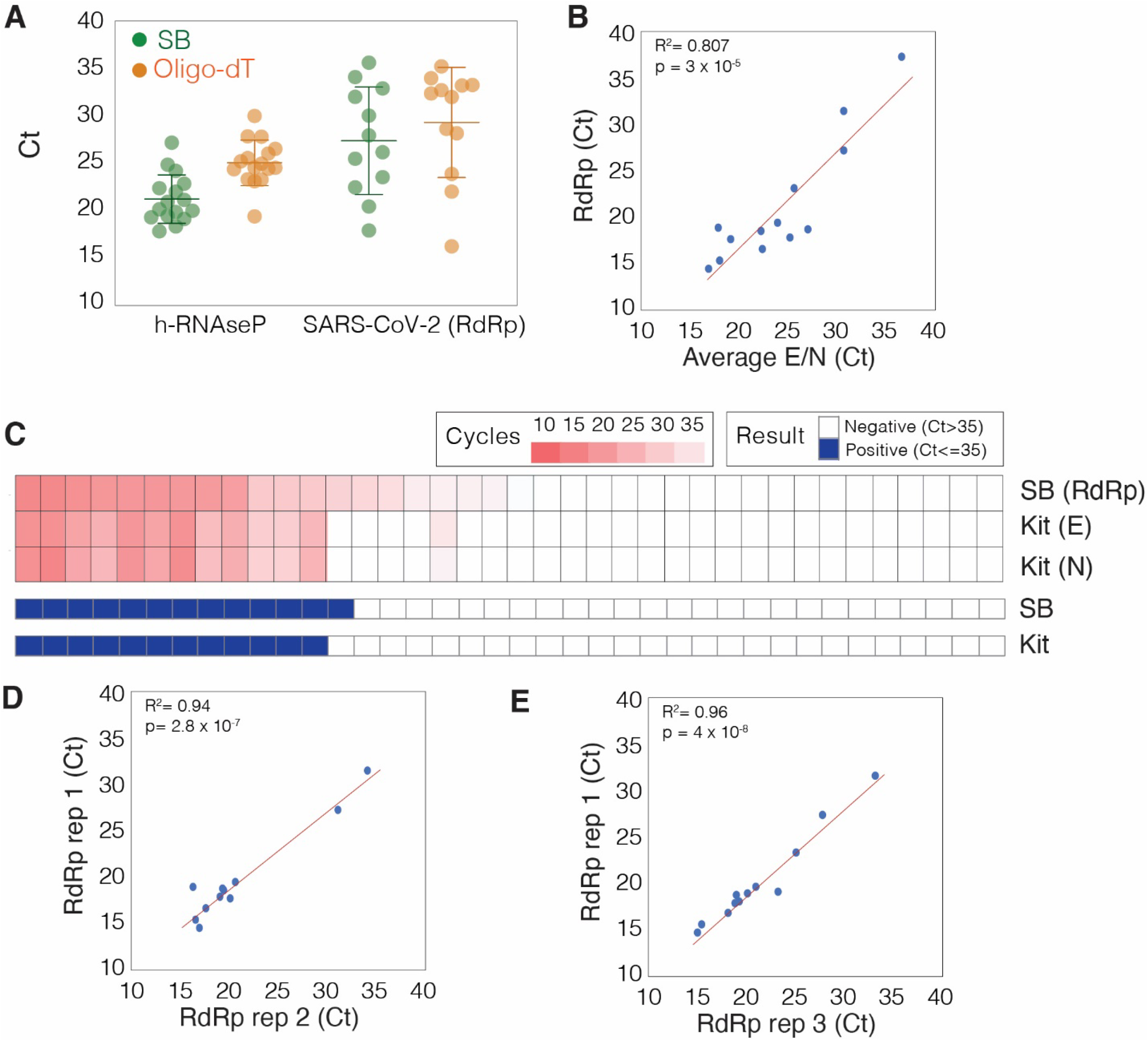
Proof of concept analysis with a series of positive and negative patients in the hospital setting. A series of patients, consecutively admitted to the emergency room of the Clínica Universidad de Navarra presenting upper airway and/or respiratory symptoms during the weeks of the COVID-19 outbreak were sampled for the presence of SARS-CoV-2 via oropharyngeal/nasopharyngeal swab. The Clinical Microbiology Service performed the standard easyMAG RNA extraction and RT-PCR using Vircell Kit. Next, the original samples were barcoded, de-identified, inactivated with Buffer RLT Plus and transported to the research center. **A**. At the research center silane bead (SB)-based and Oligo-dT-based extraction methods were tested with the SB method showing higher sensitivity. **B**. Correlation of results for all positive samples and one uncertain sample between silane beads and the standard commercial kit. **C**. Pair comparison between the two methods. **D & E**. The SB-based method obtained high reproducibility with different operators.

Next, we validated the SB method with an additional cohort of 48 clinical samples including 13 positive samples, 25 negative samples, and 10 mock controls. The new method showed 100% sensitivity (95% CI: 75·3-100%) and 96·0% specificity (95% CI: 79·6-99·9%), with a positive likelihood ratio of 25·0 (3·66-170·6) and a highly linear correlation of cycle threshold (Ct) values (R^2^=0·807 p=3E-05) when compared with the standard column-based RNA extraction method (Figure 2B) despite detecting different sets of viral genes (N/E or RdRp). One sample of 48 was identified as positive (Ct=32·2) that had not been previously diagnosed with SARS-CoV-2 with the standard method (Figure 2C). No false negatives were produced. The test was replicated three times by three different operators, demonstrating very low inter-operator variability (Figure 2D & E) (R^2^=0·94 p=1·2E-06; R^2^=0·96 p=4E-08).

In order to test the applicability of the method for comprehensive testing of vulnerable communities, 225 samples from a nursing home including 138 residents and 87 staff were obtained and processed (Figure 3A). Among residents, 63 samples produced a positive result for SARS-CoV-2 (45·7%), while 71 samples were negative (50·4%), and 4 were uncertain (2·9%). Among staff, 14 of 87 were positive (16·1%), 71 were negative (81·6%), and 2 were uncertain (2·3%) (Figure 3B). When examining the medial records of the residents, no statistical differences were found among SARS-CoV-2 negative and positive individuals, having similar prevalence of hypertension, type 2 diabetes, dyslipidemia, and similar rates of chronic neurological, cardiovascular, hepatic, or kidney diseases, and of cancer (Table 1). Only eighteen of the residents and one staff member with positive tests were mildly symptomatic (28·6% and 7·1%, respectively) at the time of sample collection.

**Table 1.** Summary of clinical characteristics

|  | **SRAS-CoV-2 Negative**<br>**N=74** | **SARS-CoV-2 Positive**<br>**N=63** | **P-Value** |
| --- | --- | --- | --- |
| Age, mean (sd) | 83.97 (9.3) | 82.5 (10.2) | 0.382 |
| Gender female, N (%) | 33 (44.6) | 26 (41.3) | 0.436 |
| Symptoms, N (%) | 9 (12.5) | 17 (27.9) | **0.030 \*** |
| Mortality 1m, N (%) | 1 (1.4) | 10 (16.1) | **0.003 \*** |
| ***Risk factors and comorbidities, N (%)*** |  |  |  |
| Hypertension | 52 (72.2) | 33 (63.5) | 0.331 |
| Type 2 Diabetes | 24 (33.3) | 11 (21.2) | 0.160 |
| Dyslipidaemia | 20 (27.8) | 17 (32.7) | 0.558 |
| Neurological disease | 35 (48.6) | 21 (40.4) | 0.465 |
| Cardiovascular disease | 22 (30.6) | 16 (30.8) | 1.000 |
| Cancer | 3 (4.2%) | 7 (13.5) | 0.093 |
| Respiratory disease | 16 (22.2) | 8 (15.4) | 0.368 |
| Kidney disease | 15 (20.8) | 7 (13.5) | 0.346 |
| Liver disease | 1 (1.4) | 4 (7.7) | 0.160 |

**Figure 3.**
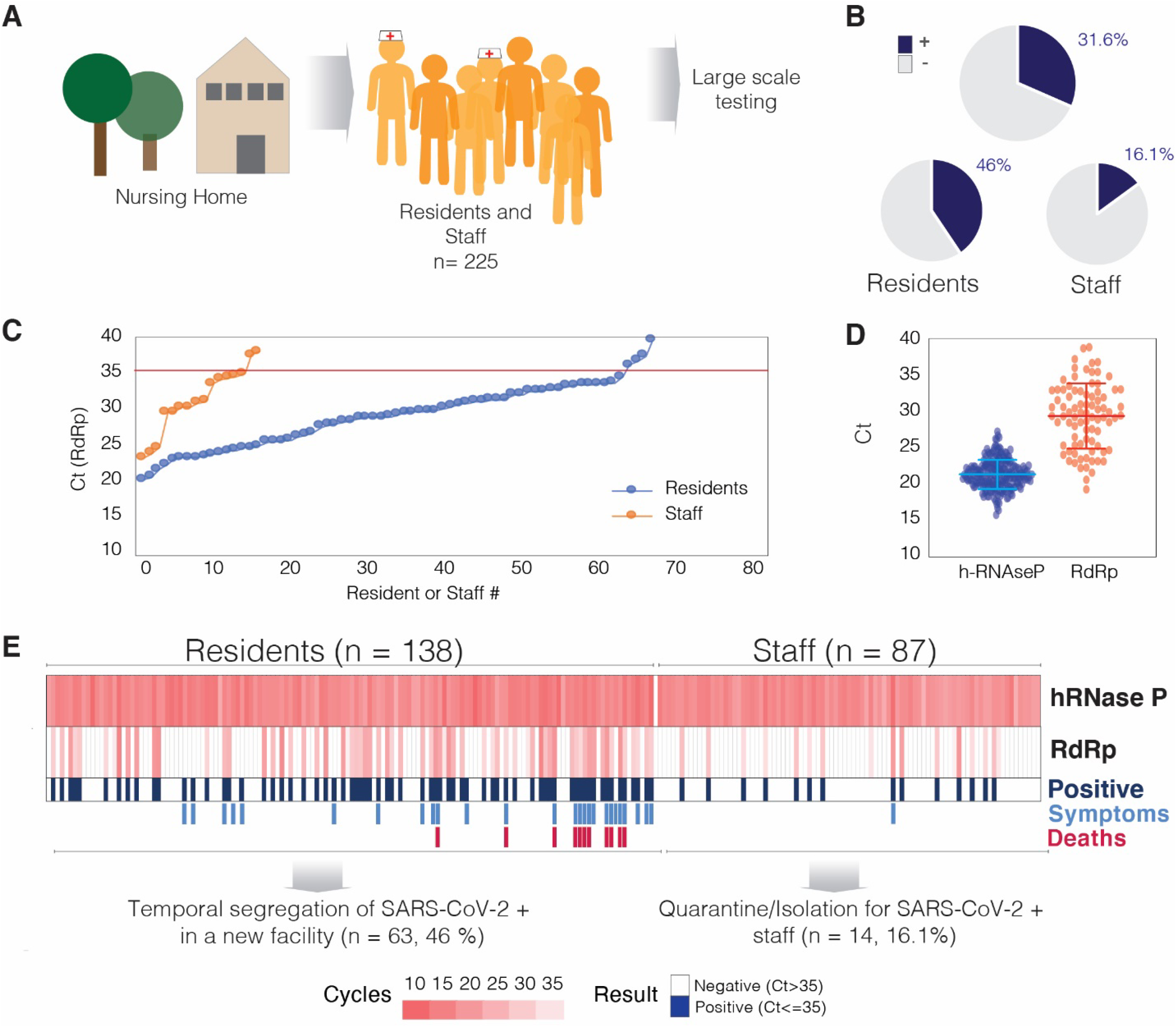
Large-scale RNA extraction and RT-PCR using silane beads allows for rapid testing of a vulnerable population. A. 137 residents and 87 staff members from a regional nursing home were sampled via oropharyngeal swab and samples were conserved in RLT Plus buffer for transport to the academic research facility. **B**. Rapid large-scale testing identified a substantial proportion of residents and staff as positive for SARS-CoV-2. **C**. Ct values (RdRp) for residents and staff show the distribution of 63 and 14 SARS-CoV-2 positives, respectively. **D**. RdRp Ct values had a wider uniform distribution across patient samples, while the internal control (h-RNaseP) was highly consistent. **E**. RT-qPCR diagnostic outcomes for residents and staff led to segregation of positive and negative residents and quarantined staff members with positive results as preventive measures.

The levels of viral detection varied among positive patients. Five samples (6·5%) had a high level of viral RNA with a Ct less than 23, and 18 samples (23·4%) had Ct between 23 and 26 (Figure 3C). Amplification of the internal control for the 225 samples was highly uniform (mean Ct = 21·21 ±2·1) in clear contrast with viral RNA variability (mean Ct = 28·8 ±5·3) (Figure 3D, p<0·001).

The results were reported to the head of the facility, who established containment protocols and established treatment decisions for each patient. SARS-CoV-2 positive staff members were put under quarantine and contacted daily for clinical follow-up. SARS-CoV-2-positive residents were segregated from SARS-CoV-2-negative residents by transfer to a different home (Figure 3E). From those negatives, 6 became symptomatic and were tested positive during follow-up. All SARS-CoV-2-negative residents were surveilled for the potential onset of new symptoms, considering previous potential contact with positive residents and staff. During one-month follow-up, 10 of the positive (15·9%) and one of the negative patients (1·3%) died.

## Discussion

All countries highly affected by COVID-19 pandemics are starting the relaxation of lockdowns of cities and regions. Since a high percentage of the population is probably not protected,^12^ a new outbreak in the next few week or months is still possible. To plan a safe transition from lockdown to regular social life, there is thus the need to rapidly identify asymptomatic viral carriers, especially within at-risk environments such as nursing homes, before the virus can be transmitted. Here we show a scalable method for large-scale RNA extraction and PCR-mediated detection of SARS-CoV-2 in pharyngeal swabs. We have shown its reliability, consistency, reproducibility, and high sensitivity. Using this methodology, we have tested all residents and the entire staff of a nursing home revealing a high percentage of asymptomatic carriers. The residents that tested positive for COVID-19 were at higher risk of short-term mortality than the negative residents. Finally, appropriate planning for adequate isolation and clinical surveillance was established for residents and health care staff.

The silane bead-based RNA extraction protocol was highly reproducible across different operators. Two key concepts for effective scaleup are multiplexing, such as through the use of multi-well plates and multichannel pipetting, and automation, such as through the use of liquid-handling robotic platforms. This workflow can be easily carried out by research laboratories, with commonly high numbers of thermocyclers and experienced personnel, in a way that does not oversaturate regular microbiology laboratories. Rapid repeat testing could provide some insight into degree of viral shedding and duration of contagion. This information could also benefit local policy makers for epidemiologic modeling of a specific region.

In addition to nursing home residents, health care workers that could act as asymptomatic spreaders were placed under quarantine. The relevance of adequately identifying these spreaders has been recently highlighted as a major factor of viral spreading in several countries affected by the pandemics.^13–15^

The protocol is cost-efficient and is not subject to the availability of commercial RNA extraction kits, bypassing the major bottleneck to PCR testing worldwide. Besides saving lives in nursing homes, this method could assist in facilitating the implementation of safe and rapid return to normal social and economic activity.

## Methods

### Patients

The assay set-up cohort included 38 patients consecutively admitted to the Emergency Room of the Clínica Universidad de Navarra (Pamplona, Spain) presenting symptoms suggestive of COVID-19. Nasopharyngeal and oropharyngeal swab specimens were collected and combined into 2 mL viral transport medium. Fourteen patients (36·8%) had a positive PCR for SARS-CoV-2 sequences. The median Ct was 24·13 for the N gene (N) and 24·18 for the E gene (E). The median age was 61, and 57·1% were women. Fever (84·6%), cough (64·29%), asthenia (50%), and dyspnea (42·86%) were the main symptoms. At baseline, 28·6% needed some degree of oxygen supplementation, and 50% had radiologic pneumonia.

To evaluate the potential of large-scale testing in real-life scenarios, 225 individuals from a nursing home were selected. Oropharyngeal swab samples were collected from all residents and staff and placed in 800 μl Buffer RLT Plus (Qiagen) and transferred to the academic research center for RNA extraction.

### Magnetic bead-based large-scale RNA extraction and RT-qPCR

RNA was extracted from swab samples of 16 de-identified patients using two procedures: (i) silane magnetic beads (SB) and (ii) oligo-dT magnetic beads (Thermofisher) following manufacturer’s instructions. A schematic summary of the SB protocol is presented in Figure 1 and has been uploaded to protocols.io (dx.doi.org/10.17504/protocols.io.bfmajk2e). Briefly, 150 μL of sample in RLT Plus buffer is transferred to 96-well plates and then mixed with previously washed silane beads in RLT Plus buffer (10 μL) and 100% ethanol (70 μL). After incubation to allow RNA binding (5 min), and after all washing steps, the beads are pelleted using a DynaMag™-96 side skirted magnet (Thermofisher) and supernatant is removed. Two washes with 75% ethanol (200 μL and 150 μL) purify the RNA, which is then eluted in 10 mM Tris pH7·5 at 37C (30-50 μL). The eluted RNA is transferred to a new 96-well plate which can be loaded manually or via robot onto a PCR plate for subsequent real-time PCR detection.

A single-step reverse transcription coupled to a real-time PCR multiplexed assay was used for the detection of SARS-CoV-2 RNA-dependent RNA Polymerase (RdRp) and human RNase P (internal control) using TaqMan probes (2019-nCoV: Real-Time Fluorescent RT-PCR kit, BGI). Positivity cut-off based on the receiver operator characteristic curve from testing clinical samples as specified by the manufacturer instructions was Ct > 38. However, during test validation several non-template negative controls generated Ct values for RdRp between 35 and 39. Therefore, Ct values below 35 cycles were deemed positive for SARS-CoV-2, while samples with Ct values between 35 and 40 were rerun. If Ct values on the rerun were between 35 and 40 for a second time, they were considered “uncertain”.

### Standard RNA extraction and RT-qPCR

RNA extraction from swabs was performed using the NUCLISENS easyMAG Kit (Biomerieux) and the identification of SARS-CoV-2 transcripts encoding N and E was performed using a commercial kit (Vircell), both according to manufacturer instructions.

### Statistics

Correlations were analyzed by Pearson’s χ^2^ test, and variance between RdRp and RNaseP Ct were analyzed using Mann-Whitney and unpaired t test, with p<0·05 considered significant.

### Ethical Statement

The study was approved by the Medical Research Ethical Committee of the University of Navarra (protocol number 2020·077). Informed consent was obtained by all participants of the study.

## Data Availability

All data needed to evaluate the conclusions in the paper are present in the paper. All other data supporting the findings of this study are available from the corresponding authors upon reasonable request.

## Author Contributions

AGS, JAR, DLA and JP conceived and designed experiments.

NDW, AGS, JAR, JPU, DA, ES, SMG, CO, LO, MNA, and DLA performed experiments.

NDW, JAR, JPU, SMG, JA, and DLA analyzed the data.

MFA, FA, FCT, MMO, RSO and JA sampled and performed test studies on patients.

NDW, JA and DLA wrote the manuscript and all authors participated in the editorial process and approved the manuscript.

## REFERENCES

1. European Centre for Disease Prevention and Control. Rapid Risk Assessment: Coronavirus disease 2019 (COVID-19) in the EU/EEA and the UK– ninth update (2020)

2. Instituo de Salud Carlos III. Situación de COVID-19 en España. https://covid19.isciii.es/ [last consult on May 10th 2020]

3. Sosa-Troya M. “Data shows over 17,500 confirmed or probable Covid-19 deaths at Spain’s care homes”. El Pais. https://english.elpais.com/society/2020-05-07/data-shows-over-17500-confirmed-or-probable-covid-19-deaths-at-spains-care-homes.html [last consult on May 10th 2020]

4. Arons MM, Hatfield KM, Reddy SC, et al. Presymptomatic SARS-CoV-2 Infections and Transmission in a Skilled Nursing Facility. N Engl J Med doi:10.1056/NEJMoa2008457

5. Kissler SM, Tedijanto C, Goldstein E, Grad YH, Lipsitch M. Projecting the transmission dynamics of SARS-CoV-2 through the postpandemic period. Science. 2020 doi:10.1126/science.abb5793.

6. He X, Lau EHY, Wu P, et al. Temporal dynamics in viral shedding and transmissibility of COVID-19. Nat Med 2020 https://doi.org/10.1038/s41591-020-0869-5

7. Whitman JD, Hiatt J, Mowery CT, et al. Test performance evaluation of SARS-CoV-2 serological assays. 2020 https://covidtestingproject.org/

8. Peto J, Alwan NA, Godfrey KM, et al. Universal weekly testing as the UK COVID-19 lockdown exit strategy. Lancet 2020 https://doi.org/10.1016/S0140-6736(20)30936-3

9. Jha AK, Tsai T, and Jacobson B. Why we need at least 500,000 tests per day to open the economy — and stay open. Globalepidemics.org https://globalepidemics.org/2020/04/18/why-we-need-500000-tests-per-day-to-open-the-economy-and-stay-open/ [last consult on May 10th 2020]

10. Bruce EA, Huang M-L, Perchetti GA, et al. Direct RT-qPCR detection of SARS-CoV-2 RNA from patient nasopharyngeal swabs without an RNA extraction step. Biorxiv.org https://www.biorxiv.org/content/10.1101/2020.03.20.001008v2 (2020)

11. Maxmen A. Untapped potential: More US labs could be providing tests for coronavirus. Nature 2020 doi:10.1038/d41586-020-01154-6

12. Garcia-Basteiro AL, Moncunil Gl, Tortajada M, et al. Seroprevalence of antibodies against SARS CoV-2 among health care workers in a large Spanish reference hospital. (2020) Medrxiv.org https://doi.org/10.1101/2020.04.27.20082289

13. Trabucchi M, De Leo D. Nursing homes or besieged castles: COVID-19 in northern Italy. Lancet Psychiatry. 2020 May;7(5):387–388. doi:10.1016/S2215-0366(20)30149-8. PubMed PMID: 32353267; PubMed Central PMCID: PMC7185948

14. McMichael TM, Currie DW, Clark S, et al. Epidemiology of Covid-19 in a Long-Term Care Facility in King County, Washington. N Engl J Med. 2020 Mar 27. doi:10.1056/NEJMoa2005412. [Epub ahead of print] PubMed PMID: 32220208; PubMed Central PMCID: PMC7121761.

15. Htun HL, Lim DW, Kyaw WM, et al. Responding to the COVID-19 outbreak in Singapore: Staff Protection and Staff Temperature and Sickness Surveillance Systems. Clin Infect Dis. 2020 Apr 21. pii: ciaa468. doi:10.1093/cid/ciaa468. [Epub ahead of print] PubMed PMID: 32315026; PubMed Central PMCID: PMC7188160.

